# CT validation of intraoperative imageless navigation (Naviswiss) for component positioning accuracy in primary total hip arthroplasty in supine patient position: A prospective observational cohort study in a single-surgeon practice

**DOI:** 10.1101/2023.07.10.23292202

**Authors:** Corey Scholes, Tobias Schwagli, John Ireland

**Affiliations:** EBM Analytics, Sydney, NSW, Australia; Medivation, Brugg, Switzerland; Sydney Bone and Joint Clinic, Sydney, NSW, Australia

## Abstract

The aim of this study was to report on the validity of the Naviswiss handheld image-free navigation device for accurate measurement of THA component positioning intraoperatively, in comparison with the three-dimensional (3D) reconstruction of computed tomography (CT) images as gold standard.

**Methods:** A series of patients presenting to a single-surgeon clinic with end-stage hip osteoarthritis received primary hip arthroplasty with anterolateral muscle-sparing surgical approach in the supine position. Imageless navigation was applied during the procedure with bone-mounted trackers applied to the greater trochanter and ASIS. Patients underwent routine CT scans before and after surgery and these were analysed using three-dimensional reconstruction to generate cup orientation, offset and leg length changes which were compared to the intraoperative measurements provided by the navigation system. Estimates of agreement between the intraoperative and image-derived measurements were assessed with and without correction for bias and declared cases with potential measurement issues.

**Results:** The mean difference between intraoperative and postoperative CT measurements was within 2° for angular measurements and 2mm for leg length. Absolute differences for the two indices were within 5° and 4mm. Mean bias was 1.9 - 3.6° underestimation for cup orientation and up to 2mm overestimation for leg length change, but absolute thresholds of 10° and 10mm were not exceeded by 95% limits of agreement (LOA), especially after correction for bias. Four cases (12%) were declared intraoperatively for issues with fixation on the greater trochanter. Inclusion of these cases generated acceptable accuracy overall and their omission failed to improve between-case variability in accuracy or LOA for both offset and leg length.

**Conclusions:** The accuracy of the Naviswiss system applied during primary THA in supine patient position and anterolateral surgical approach falls within clinically acceptable recommendations for acetabular cup placement, femoral offset and length length. With refinements to surgical technique to adapt to the navigation hardware, the system could be further improved with regression-based bias correction.

## Introduction

The number of primary total hip arthroplasties (THA) in Australia has risen 125% since 2003, with revision burden reported as 7.6% at the end of 2021 (Australian Orthopaedic Association National Joint Replacement Registry (AOANJRR), n.d.). Accurate alignment of the implant components and limb length equalisation during THA are essential for minimising the risk of revision surgery (Migliorini et al., 2022). Poor positioning of the acetabular cup can cause dislocation, impingement and instability, while a large leg length discrepancy increases the risk of back pain, gait impairment and overall patient dissatisfaction (Domb et al., 2015; Liebs et al., 2014; Renkawitz et al., 2016; Tanaka et al., 2010) .

Robotic-assisted THA can improve the accuracy of implant placement and significantly reduce leg length discrepancies (Kumar et al., 2021). However, the surgical time is prolonged and there are no significant improvements in the rate of complications and implant survivorship (Kumar et al., 2021; Kunze et al., 2022; Singh et al., 2021). The financial investment and time required to adopt these systems have also prevented their widespread use (Renner et al., 2017). Portable navigation systems have been developed to offer a cost-effective, user-friendly and minimally invasive solution, and have been demonstrated to improve component positioning (Shigemura et al., 2021) even when used with different surgical approaches (Bradley et al., 2019; Grosso et al., 2016; Hayashi et al., 2019).

The Naviswiss (Naviswiss AG, Brugg, Switzerland) is a portable imageless navigation device equipped with an infrared stereo camera and an inertial measurement unit to facilitate implant positioning intraoperatively. The accuracy of the system has been reported with a <3° mean absolute error for cup inclination and anteversion when tested in the supine position via an anterolateral THA approach (Hasegawa et al., 2022), and <3.5° for cup orientation using a direct anterior THA approach with fluoroscopy (Ong et al., 2022). However, clinical data for the system has not been reported in Australia, and there is a dearth of information regarding femoral offset and leg length discrepancy.

As such, a trial protocol was developed to assess the accuracy of the Naviswiss system in measuring acetabular cup inclination, acetabular cup version, femoral offset and leg length discrepancy (Ektas et al., 2020). The aim of this study was to report on the validity of the Naviswiss handheld image-free navigation device for accurate measurement of THA component positioning intraoperatively, in comparison with the three-dimensional (3D) reconstruction of computed tomography (CT) images as gold standard.

## Methods

### Patient Selection

The study was embedded within a prospective observational clinical registry, for which ethical approval was obtained from a National Health and Medical Research Council certified human research and ethics committee (HREC) (Bellberry; HREC 2017-07-499). The study was also registered with the Australian New Zealand Clinical Trials Registry (ACTRN12618000317291). Adult patients (>18 years) were included in the study if they presented to the participating surgeon with end-stage or rheumatoid arthritis and elected to undergo THA.

Patients who were unable to provide informed consent, or had declined or revoked consent were excluded from the study. Patients were additionally excluded if: they had severe contralateral hip deformity or dysplasia; required a simultaneous bilateral procedure; required an ipsilateral revision procedure; had a short-stem component implanted; were lost to follow-up; use of the navigation system was completely abandoned; received a posterior surgical approach; were revised prior to post-operative imaging being performed; where hardware was unavailable or failed such that intraoperative data could not be retrieved.

### Surgical Technique

Patients were administered preoperative antibiotic prophylaxis and were placed in supine position on a radiolucent table to allow intraoperative imaging of cup and stem positions. Once patients were prepped and draped, the iliac crest was identified and a tracker fixed prior to registration of anatomical landmarks. An Anterolateral muscle sparing approach was utilised to expose the hip joint and trochanteric region. A screw with a serrated washer was inserted into the lateral trochanter near the caudal attachment of the Gluteus medius, and a stalked tracker inserted to clear the soft tissues. Registration of the hip joint was then performed. The final intra-operative component positions were logged by the navigation system and transferred by electronic form for further analysisAll surgeries were performed by the senior author.

### Measurement of component positioning

Patients underwent a postoperative CT at the 6 week follow up, which was retrieved and analysed. The primary study outcomes were extracted for analysis as previously described (Ektas et al., 2020; Scholes et al., 2023)

● Acetabular cup inclination (ACI) - Angle between the acetabular and longitudinal axes when projected onto the functional pelvic plane (FPP)
● Acetabular cup version (ACV) - Angle between the acetabular axis and the FPP
● Femoral offset (FO) - Difference between the hip centre of rotation of the operated joint relative to its starting position at the initial assessment in the coronal plane (medial-lateral) within the pelvic coordinate system
● Leg length discrepancy (LLD) - Change in the distance between the greater trochanter tag and the hip centre of rotation added to the change in the distance between the centre of the acetabulum and the centre of the cup in the transverse plane (superior-inferior)

CT images were obtained pre- and postoperatively in DICOM (Digital Imaging and Communications in Medicine) format, and information relating to the diagnosis, study or surgery was removed for blinded analysis by an independent researcher.

Inclination and version of the acetabular cup was measured on DICOM files using an online software (3D Slicer, www.slicer.org) (Fedorov et al., 2012) as previously described (Ektas et al., 2020; Scholes et al., 2023). FO and LLD were measured through assessment of anatomical landmarks picked in pre- and postoperative scans. Coordinate systems for the pelvis and femur were determined based on the anatomic landmarks for the postoperative CT assessments. Parameters were expressed relative to the FPP, with the origin placed at the centre of the line connecting the left and right ASIS.The postoperative position of the cup centre was compared with the native hip COR determined from the preoperative CT. FO and LLD was reported as the pre-to-post change of the femoral coordinate frame relative to pelvis FPP coordinates in the coronal (mediolateral) and transverse (inferior-superior) planes respectively.

## Data and Statistical Analysis

### Missing data

Missing data was identified predominantly from intraoperative measurements where technical issues precluded retrieval of certain measurements within a case, but not all (exclusion criterion). Due to the low proportion of missing data <10%, case-wise deletion was performed to restrict analysis on each outcome measure .

### Reliability

Intraobserver reliability for the image-based measures, including inclination and anteversion relative to the table orientation as well as cup position changes, was assessed using intraclass correlations, with a two-way random effects model on a sample of nine cases. Case identifier and observation (intra-observer) or observer (inter-observer) were considered the random effects. The ratings were performed by the same observer on three occasions separated by a minimum of 1 week, with blinding to previous measures as well as measures by other observers and identified only by case identifier. Standard error of measurement was defined by the root mean square error from the repeated measures analysis of variance <u>(Weir and Vincent 2020)</u> with observation or observer as the repeated factor. Standard error of measurement describes the average deviation from one measurement to the next that can be attributed to random observer error.

Interobserver reliability for the image based measures, was derived from the median of the observations from the primary observer as well as singular observations from two additional observers who performed their measurements independently on the same sub-sample (N = 9). Intraclass correlation with two way mixed effects models were also calculated, and typical error measurement was calculated in the same manner as for interobserver reliability.

### Agreement and Bias Assessment (Correction)

The calculation of agreement and bias assessment replicates the method described in Scholes et al 2023, and is summarised here for clarity. Mean deviation (delta) for each measure was calculated and summary statistics generated using a bootstrapping approach and repeated with delta converted to absolute values. Bland_altman plots were generated and limits of agreement calculated with a previously published formula (Bland et al., 1999; Giavarina, 2015) Bias was assessed using linear regression on each measure with adjustments for age, BMI and sex. Bias-corrected estimates of each measure were generated from regression model predictions. Alternative correction for offset and leg length change was performed by dropping values where an intraoperative declaration was made and the summary agreement analysis repeated as described above (Supplementary material 1). All statistical analyses were performed using Stata (v17.1, Statacorp, USA), with alpha set at 5% to indicate significant effects where appropriate.

## Results

### Patient characteristics

A consecutive sample of 54 primary cases were assessed for eligibility with 34 included for analysis (Figure 1). The analysis cohort comprised 42% females, had a mean age at surgery of 60.8 years (IQR 50.3 - 70) and a mean body mass index (BMI) of 29.6 (IQR 27.3 - 32.4).

**Figure 1:**
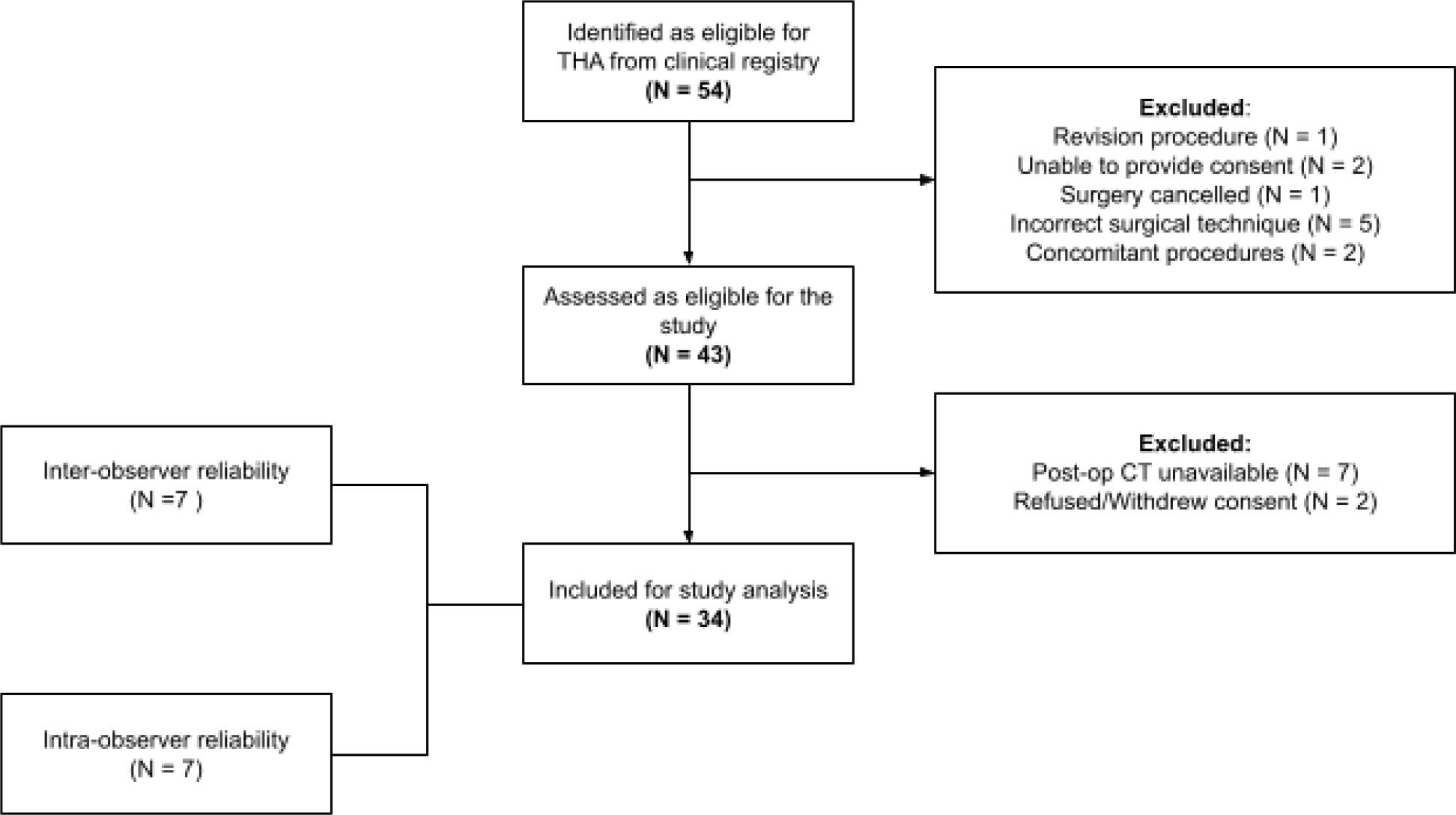
STROBE diagram (von Elm et al., 2007) of patient inclusion into the study analysis

### Imaging Reliability

The imaging analysis demonstrated adequate reliability for both within and between observers for the measures of interest (Tables 1 and 2), with intraobserver standard error of measurement ranging from<1° for cup anteversion and inclination and <1.1mm for femoral offset and leg length changes (Table 3).

**Table 1:** Individual and average intra-class correlations for **intra**observer, image-based measurements

|  | Individual |  |  | Average |  |  |
| --- | --- | --- | --- | --- | --- | --- |
|  | ICC | 95%LCI | 95%UCI | ICC | 95%LCI | 95%UCI |
| Inclination_ APP | 0.99 | 0.97 | 1.00 | 0.997 | 0.99 | 1 |
| Inclination_ FPP | 0.986 | 0.95 | 1.00 | 0.995 | 0.98 | 1 |
| Version_ APP | 0.996 | 0.986 | 1.00 | 0.998 | 0.99 | 1 |
| Version_ FPP | 0.996 | 0.987 | 1.00 | 0.998 | 0.996 | 1 |
| Offset | **0.868** | 0.62 | 0.97 | 0.95 | 0.83 | 0.99 |
| LLD | 0.987 | 0.955 | 1.00 | 0.995 | 0.984 | 1 |

**Table 2:** Individual and average intra-class correlations for **inter**observer, image-based measurements

|  | Individual |  |  | Average |  |  |
| --- | --- | --- | --- | --- | --- | --- |
|  | ICC | 95%LCI | 95%UCI | ICC | 95%LCI | 95%UCI |
| Inclination_ APP | 0.97 | 0.87 | 1.000 | 0.99 | 0.95 | 1 |
| Inclination_ FPP | 0.96 | 0.86 | 0.990 | 0.987 | 0.948 | 1 |
| Version_ APP | 0.98 | 0.94 | 1.000 | 0.99 | 0.978 | 1 |
| Version_ FPP | 0.988 | 0.959 | 0.988 | 0.996 | 0.986 | 1 |
| Offset | **0.866** | 0.604 | 0.973 | 0.951 | 0.821 | 0.99 |
| LLD | 0.945 | 0.823 | 0.989 | 0.98 | 0.933 | 0.996 |

**Table 3:** Standard error of measurement for image-based analysis of cup position and orientation

|  | Intraobserver | Interobserver |
| --- | --- | --- |
| Inclination_ APP (deg) | 0.63 | 0.82 |
| Inclination_ FPP (deg) | 0.57 | 0.82 |
| Version_ APP (deg) | 0.62 | 0.90 |
| Version_ FPP (deg) | 0.55 | 0.76 |
| Offset (mm) | 1.08 | 1.08 |
| LLD (mm) | 0.73 | 1.05 |

### Agreement - Intraoperative to Imaging

#### Thresholds and declared observations

Intraoperative declarations were made for four patients, with loss of fixation of the tracker on the greater trochanter (n = 4). A number of patients were observed to exceed the specified measurement thresholds and had no intraoperative declarations (Table 4). Two patients included in the analysis had an intraoperative declaration recorded, however their measurements did not exceed the threshold boundaries.

**Table 4:** Patients without an intraoperative declaration that exceeded the measurement <u>thresholds</u>

| Measurement | Threshold | Units | Cases above threshold | Proportion above threshold |
| --- | --- | --- | --- | --- |
| Threshold - Inclination | 10 | ° | 2 | 5.9% |
| Threshold - Version | 10 | ° | 2 | 5.9% |
| Threshold - Offset | 10 | mm | 1 | 2.9% |
| Threshold - LLD | 10 | mm | 1 | 2.9% |

The mean delta (bias) between the intraoperative measurements and the postoperative imaging are summarised in (Table 5). The 95% limits of agreement for uncorrected data exceeded 10 degrees for inclination and version, and 10mm for offset and LLD respectively (Table 5, Figure 2). The linear fit of the average to the delta indicated that the bias between the navigation and the CT measurements was not constant across magnitude for inclination, version and offset (Figure 2, Table 6). Overall, 91.2% of cases (95%CI 76.3 - 98.1) were within 10° of the image-measured for both inclination and version (Figure 3).

**Figure 2:**
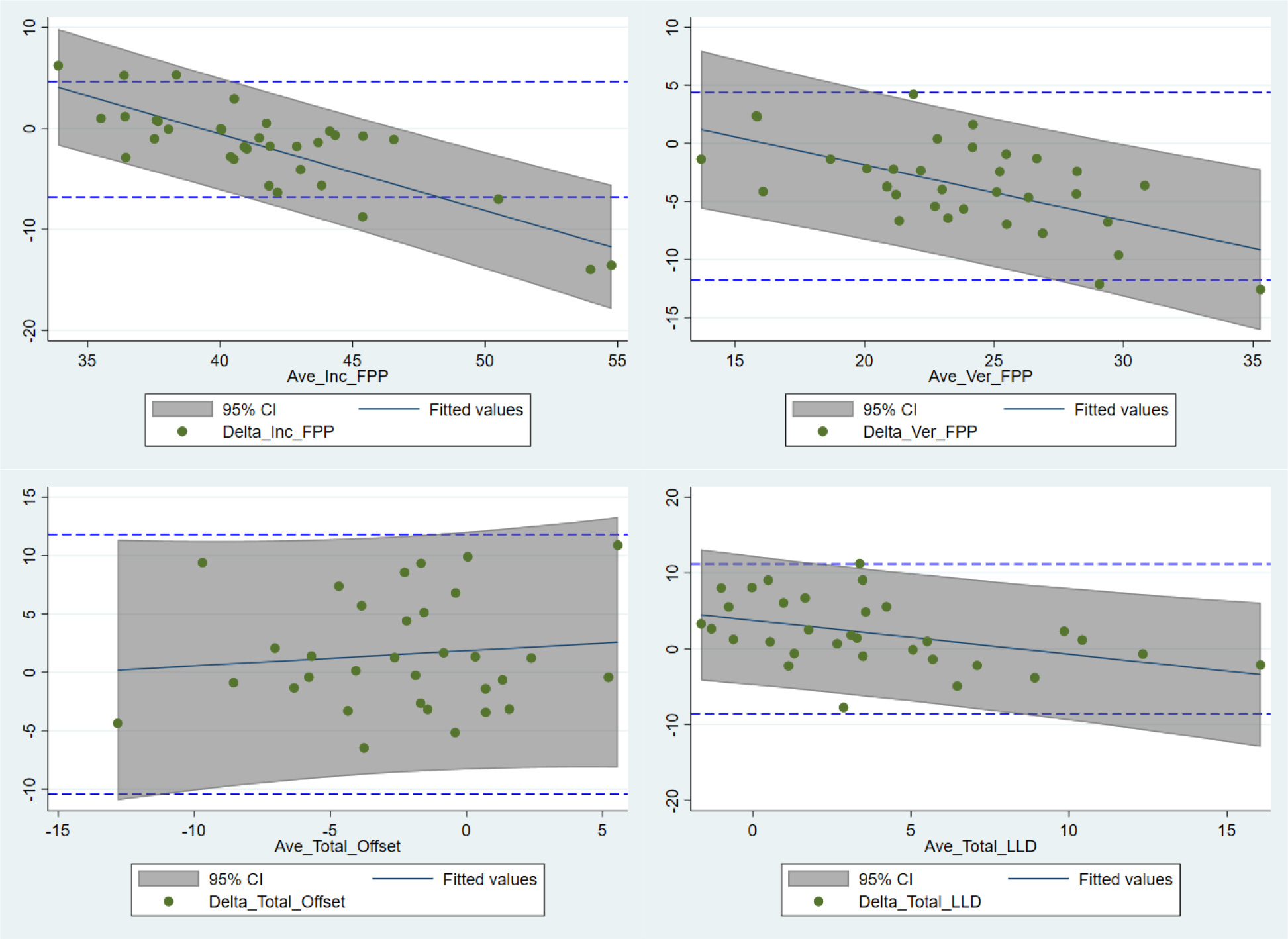
Bland-Altman plots with 95% limits of agreement for inclination,version, offset and leg length. Regression fits with shaded areas denoting 95% prediction intervals indicate the relationship between magnitude and agreement.

**Figure 3:**
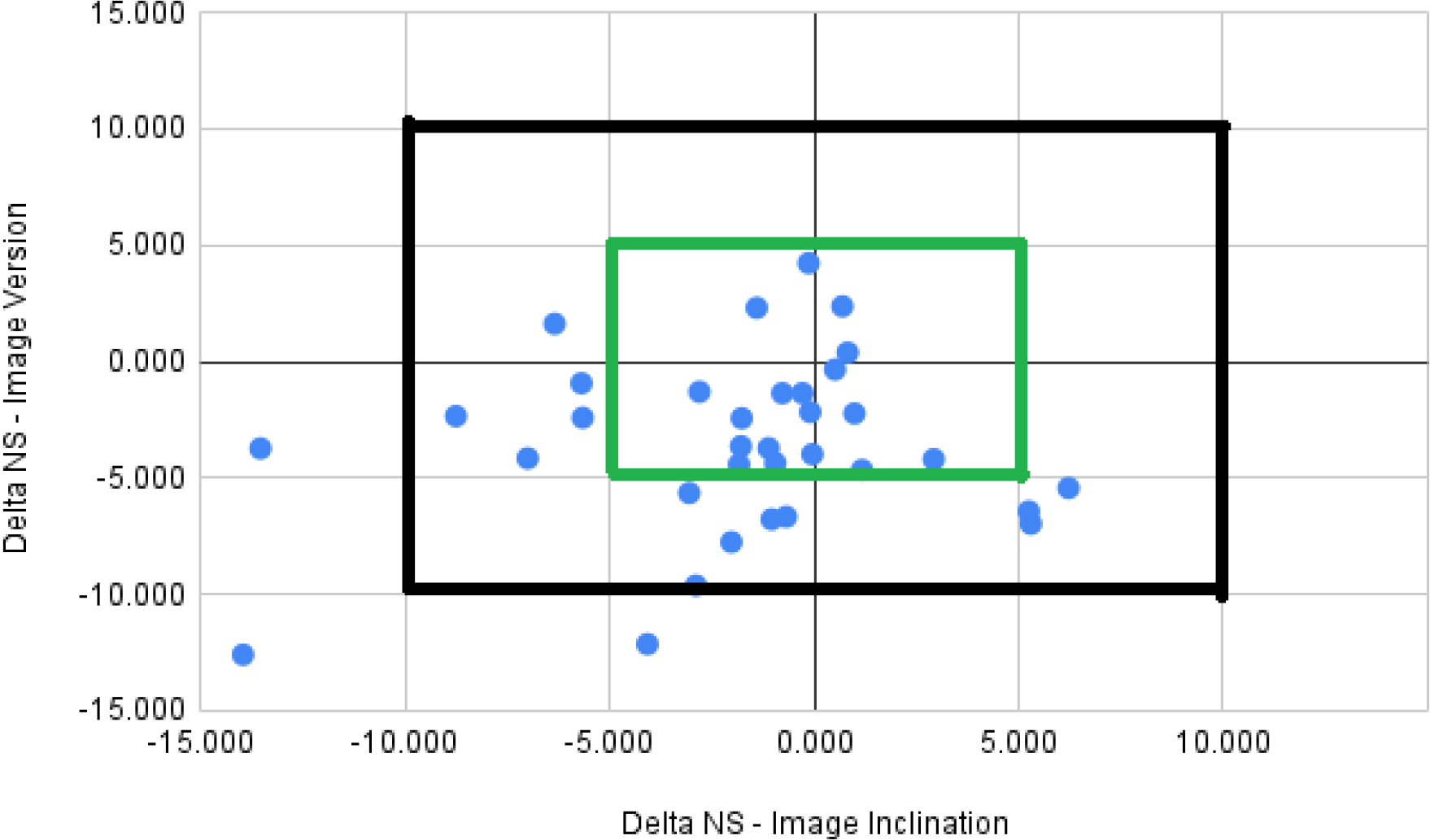
Scatterplot of delta in version versus inclination for all cases. Outer box - 10° threshold; Inner box - 5° threshold.

**Table 5:**
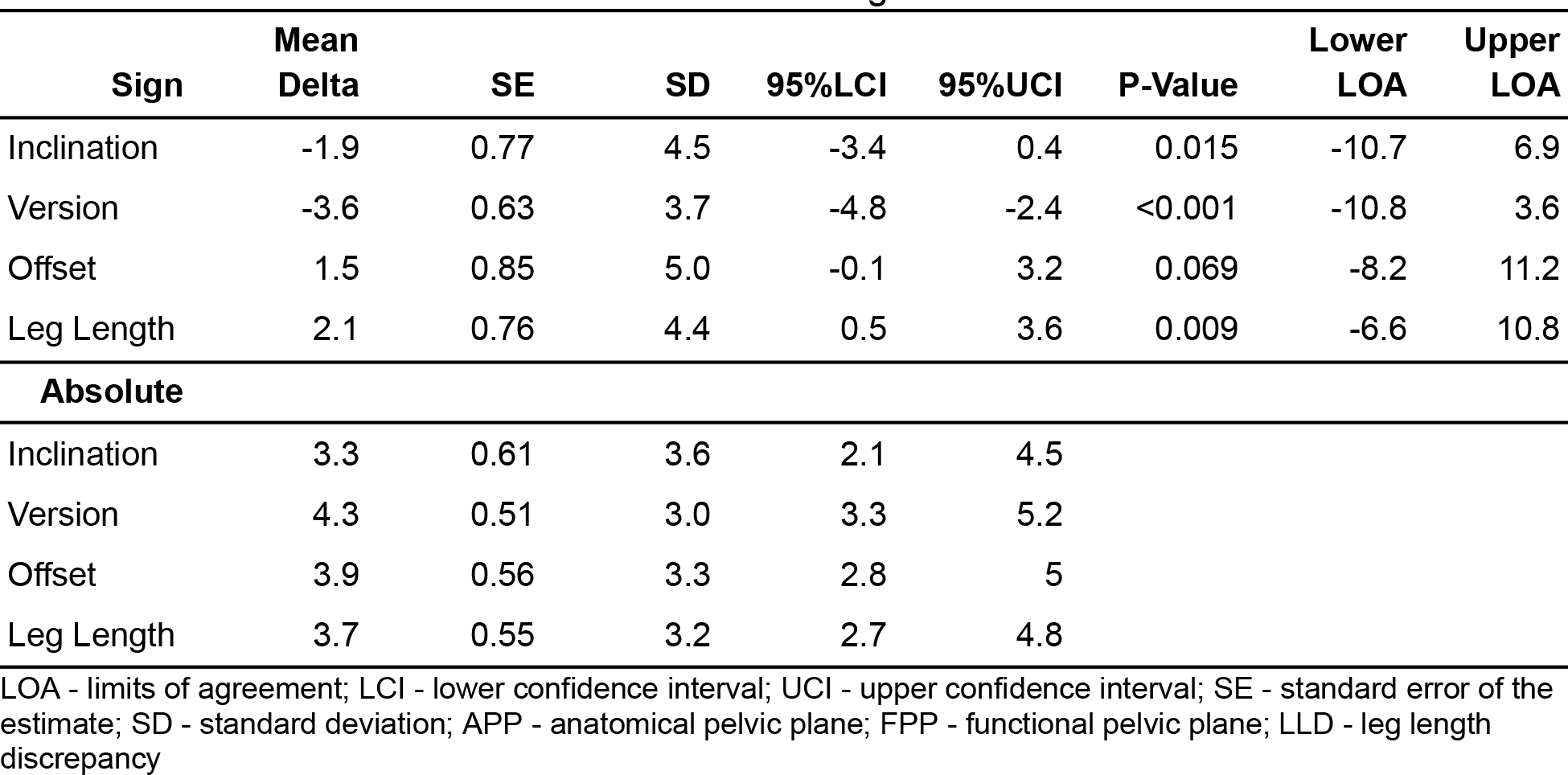
Summary of mean differences between intraoperative and image-based measurements. P-value is relative to mean delta being different to zero

| **Sign** | **Mean Delta** | **SE** | **SD** | **95%LCI** | **95%UCI** | **P-Value** | **Lower LOA** | **Upper LOA** |
| --- | --- | --- | --- | --- | --- | --- | --- | --- |
| Inclination | -1.9 | 0.77 | 4.5 | -3.4 | 0.4 | 0.015 | -10.7 | 6.9 |
| Version | -3.6 | 0.63 | 3.7 | -4.8 | -2.4 | <0.001 | -10.8 | 3.6 |
| Offset | 1.5 | 0.85 | 5.0 | -0.1 | 3.2 | 0.069 | -8.2 | 11.2 |
| Leg Length | 2.1 | 0.76 | 4.4 | 0.5 | 3.6 | 0.009 | -6.6 | 10.8 |
| **Absolute** |  |  |  |  |  |  |  |  |
| Inclination | 3.3 | 0.61 | 3.6 | 2.1 | 4.5 |  |  |  |
| Version | 4.3 | 0.51 | 3.0 | 3.3 | 5.2 |  |  |  |
| Offset | 3.9 | 0.56 | 3.3 | 2.8 | 5 |  |  |  |
| Leg Length | 3.7 | 0.55 | 3.2 | 2.7 | 4.8 |  |  |  |

LOA - limits of agreement; LCI - lower confidence interval; UCI - upper confidence interval; SE - standard error of the estimate; SD - standard deviation; APP - anatomical pelvic plane; FPP - functional pelvic plane; LLD - leg length discrepancy

**Table 6:** Linear fit of average of measurements to delta of measurements

|  | **Coefficient** | **SE** | **95%LCI** | **95%UCI** | **P-Value** | **Adjusted R-sq** |
| --- | --- | --- | --- | --- | --- | --- |
| Inclination | -0.76 | 0.12 | -1.00 | -0.51 | <0.001 | 0.65 |
| Version | -0.48 | 0.12 | -0.71 | -0.25 | <0.001 | 0.35 |
| Offset | 0.13 | 0.26 | 0.38 | 0.64 | 0.617 | -0.02 |
| Leg Length | -0.45 | 0.14 | -0.72 | -0.17 | 0.001 | 0.18 |

### Factors associated with agreement and Bias Correction

The regression results indicated a significant magnitude-dependent bias for inclination (P = 0.014) and offset (P = 0.002) (Supplementary 2). Bias correction applied to the intraoperative measures removed overall bias and shrank the between-case variation (SD) of delta by 1 - 20%, and by 22 - 41% for absolute values (Table 7). Bias correction also shrank the mean absolute delta by 5 - 35% relative to the uncorrected values. Conversely, by omitting declared observations for offset and leg length (Table 8), mean absolute error was not reduced and between case variability increased by 5-7%.

**Table 7:**
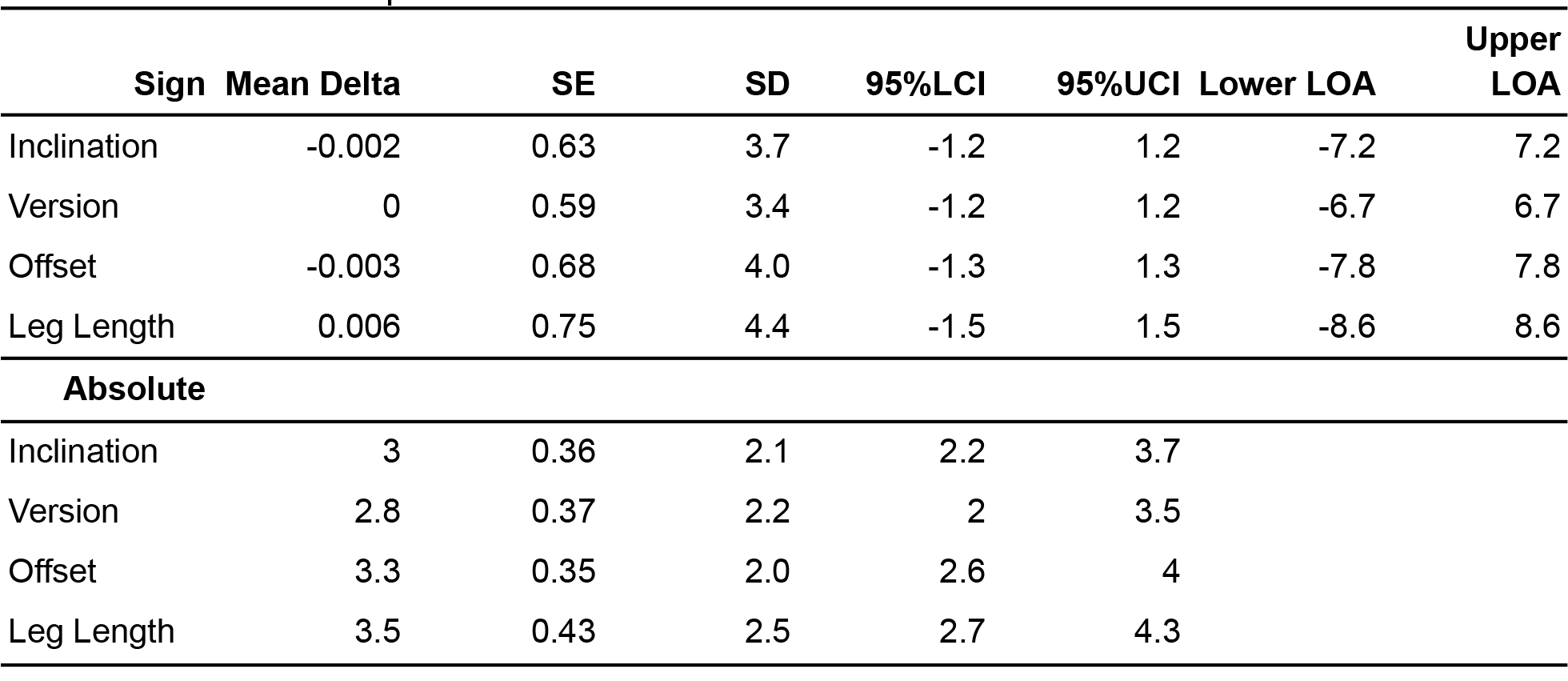
Summary of mean differences between intraoperative and image-based measurements for bias corrected intraoperative measures

|  | **Sign** | **Mean Delta** | **SE** | **SD** | **95%LCI** | **95%UCI** | **Lower LOA** | **Upper LOA** |
| --- | --- | --- | --- | --- | --- | --- | --- | --- |
| Inclination |  | -0.002 | 0.63 | 3.7 | -1.2 | 1.2 | -7.2 | 7.2 |
| Version |  | 0 | 0.59 | 3.4 | -1.2 | 1.2 | -6.7 | 6.7 |
| Offset |  | -0.003 | 0.68 | 4.0 | -1.3 | 1.3 | -7.8 | 7.8 |
| Leg Length |  | 0.006 | 0.75 | 4.4 | -1.5 | 1.5 | -8.6 | 8.6 |
| **Absolute** |  |  |  |  |  |  |  |  |
| Inclination |  | 3 | 0.36 | 2.1 | 2.2 | 3.7 |  |  |
| Version |  | 2.8 | 0.37 | 2.2 | 2 | 3.5 |  |  |
| Offset |  | 3.3 | 0.35 | 2.0 | 2.6 | 4 |  |  |
| Leg Length |  | 3.5 | 0.43 | 2.5 | 2.7 | 4.3 |  |  |

**Table 8:** Summary of mean differences between intraoperative and image-based measurements with declaration cases omitted (N = 30)

|  | **Sign** | **Mean Delta** | **SE** | **SD** | **95%LCI** | **95%UCI** | **P-Value** | **Lower LOA** | **Upper LOA** |
| --- | --- | --- | --- | --- | --- | --- | --- | --- | --- |
| Offset |  | 1.3 | 0.88 | 5.1 | -0.4 | 3 | 0.146 | -8.8 | 11.4 |
| Leg Length |  | 2.4 | 0.78 | 4.5 | 0.9 | 3.9 | 0.002 | -6.5 | 11.3 |
| **Absolute** |  |  |  |  |  |  |  |  |  |
| Offset |  | 3.8 | 0.55 | 3.2 | 2.7 | 4.8 |  |  |  |
| Leg Length |  | 3.8 | 0.59 | 3.4 | 2.6 | 4.9 |  |  |  |

## Discussion

The aim of this study was to report on the validity of an imageless navigation system (Naviswiss) for measurement of THA component positioning intraoperatively, in comparison with the three-dimensional (3D) reconstruction of computed tomography (CT) images as gold standard. The results identified reasonable accuracy for the metrics of interest with opportunities for further development identified.

The mean absolute deviation of acetabular inclination (3.3, 95% CI 2.1 - 4.5) between the navigation system and the CT-based analysis was comparable to the deviation reported by (Hasegawa et al., 2022) (2.8, 2.3 - 3.3) in the supine position, as well as Liu et al 2023 in the lateral decubitus position (3.6, 2.6 - 4.7) but higher than the pooled deviation (2.6, 2.4 - 2.8) for previous studies in the supine patient position using CT-based, imageless and accelerometry systems (Supplementary material 3 - summary of validation findings). In contrast, anteversion mean absolute deviation (4.3, 3.3 - 5.2) was greater than Hasegawa (2.8, 2.3 - 3.3), but comparable to pooled deviation for previous studies (3.6, 3.4 - 3.8). Overall, mean bias was 1.9-3.6° underestimation for cup orientation and up to 4mm overestimation for leg length change, with 95% LOA on or below 11° for orientation and 6-11mm for offset/leg length change. Absolute thresholds of 10° and 10mm were established a-priori in the study protocol (Ektas et al., 2020) and were exceeded by 95% LOA, but this was reduced by bias correction to <10. Between-patient variation in published guidelines for cup orientation vary between 5 and 12° for inclination and up to 18° for version (Harrison et al., 2014). In general, less than 10mm of LLD is considered acceptable after THA (McWilliams et al., 2013). In addition, a simulation study (Shoji et al., 2018) reported impingement and loss of motion range with a 4mm medialisation/lateralisation of the cup, although this amount of change was not justified in their methods. The bias-corrected LOA in the present study suggests that 95% of the patient sample would fall within these tolerances for cup orientation, but not for offset. The findings are comparable to Liu et al 2023, which performed the same analysis with patients in the lateral decubitus position. Better understanding of the factors contributing to error in patients, the extremes of anthropometry and morphology, irrespective of surgical approach or patient position, may provide an important direction for further research.

In any validation study, a desire to explain deviations from true agreement is a natural progression of the analysis. In this study, a biphasic pattern of magnitude-dependent bias was observed for inclination (FPP), version and leg length change, which was also noted in Liu et al 2023. The navigation system tended to overestimate at smaller average measurements and underestimate at larger averages (Figure 2). While bias correction was able to re-centre the sample around zero and reduce between-patient variation, further work is needed to validate regression-based bias correction algorithms to mitigate the magnitude-dependency (slope) (Liu et al 2023). In addition, further work may be required to improve tracker fixation for offset and leg length with 4 cases (11.8%) declared intraoperatively for issues with fixation on the greater trochanter. In contrast to Liu et al, inclusion of these cases generated acceptable accuracy overall, while their omission worsened the between-case variability in accuracy and increased the LOA for both offset and leg length. A previous study (Hasegawa et al., 2022), mentioned the potential vulnerability of the system to pin fixation on the iliac crest.

In the context of magnitude-dependent bias of the system, the most realistic explanation for deviations from true agreement are a combination of imaging measurement error (all measures), dampening of the intraoperative measurements from a combination of soft tissue coverage and draping across key anatomical landmarks, as well as soft-tissue interference due to the proximity of the surgical incision (offset, leg length). Three-dimensional CT-based measurements are considered the gold standard when determining acetabular cup position and leg length postoperatively (Bayraktar et al., 2017; Kjellberg et al., 2012; Sariali et al., 2021). The present study however, demonstrates a lower reliability of the interobserver imaging analysis with up to 1° for cup anteversion to 1.1mm for LLD. In CT-based texture analysis, it has been demonstrated that the robustness, repeatability and reproducibility of measurements is sensitive to the scanner and scanning parameters (Varghese et al., 2019). The impact of radiology environment and context on postoperative measurements in THA has not been widely explored, but cannot be disregarded when comparing the results of different investigators and investigations. In addition, the selection of manual landmarks on CT datasets, while considered reliable (ref), can still result in THA acetabular abduction and anteversion angle between-case variance of up to 2.5° (Lubovsky et al., 2010). In this study, the combination of scans performed on different scanners (patient access and convenience) and intra/inter observer error may have contributed to the typical error of the imaging measurements. For example, the typical error for offset measurement was equal to 29% of the navigation absolute error. The present results, as well as others that have relied on CT/radiograph validation, should be interpreted with these findings in mind.

While the use of imageless navigation in the supine patient position is not as vulnerable to position changes as lateral decubitus position (Liu et al 2023), many of the issues raised by Liu et al also apply to validation attempts in the supine position. Certainly, the mean BMI of the present sample are comparable to their study, while general approaches to draping and soft tissue distribution across key landmarks are almost identical. The replication of magnitude-dependent bias, as well as improved overall accuracy through bias-correction in both studies suggests that intraoperative accuracy could be further improved by targeting potential measurement dampening. Anecdotally, these issues are exacerbated with increases in patient size and the amount of soft tissue distributed around the surgical field. A potential for interference between soft tissue and the tracker fixed to the greater trochanter may be somewhat specific to the present study however, due to the location of the anterolateral surgical incision. The incision creates a segment of tissue flap that had a propensity to physically interact with the trochanter tracker without mitigation strategies in place. When combined with higher BMI patients with a greater propensity for soft tissue distribution around this area, the tissue flap could cause unpredictable deviations in measurement through the course of the procedure. Of note was the attempts at mitigation that were implemented over the course of the trial by adjusting the incision location to reduce the amount of loose tissue, as well taking additional steps to secure the flap and prevent abutment against the tracker itself. Further work is required to quantify the efficacy of these strategies.

The present study establishes that the system of interest is capable of providing comparable accuracy to similar systems in a broad range of patient populations, while identifying potential avenues for further development to achieve superior accuracy. Nevertheless, the findings and interpretations should be considered in light of the study limitations. Firstly, as discussed, variability in the imaging situation from one patient to another may have contributed to some of the errors observed in the imaging analysis. Clinical imaging in our region is referral-based, for the patient to organise a booking with a provider based on convenience and usually geographic location relative to their residence. While every attempt to standardise the imaging protocol was made with usual providers, some patients undertook their imaging studies outside of this network. Despite the mitigation attempted, there is still the potential for variable imaging parameters due to different machines being used. Secondly, comparisons with the related literature remains problematic, as introduced by Liu et al, there are a range of different analytical techniques that have been used to describe the validity of the system intraoperatively, across a broad range of population types. However, most problematic is the metric(s) used to report accuracy, which tend toward a summary of average deviation. Considering the findings of the present study, the assumption that error does not vary based on measurement magnitude clearly does not hold, although it is impossible to determine if this phenomenon has been observed elsewhere, as no other study has reported Bland-Altman plots or similar techniques to describe this relationship. This makes an average deviation inappropriate to describe average error and future work should report using regression-based metrics. Thirdly, while this study does not represent the learning curve for the operating surgeon, the early part of the present series was characterised by continued development of the system with the manufacturer, with a limited number of cases having to be abandoned entirely due to technical faults (e.g system lockup, dropped tracker tags). This may have inserted a low level of selection bias into the series and may be a consideration for clinical interpretation for those less experienced with imageless navigation in THA. Lastly, the current series has been deliberately restricted to primary, relatively uncomplicated THA, which may limit its generalisability to more extreme morphological presentations (revision, tumour) where navigation may of specific benefit. Further work is required to extend the validity of the system to this broader case-mix.

## Conclusion

The navigation system assessed in a primary THA cohort for end-stage hip osteoarthritis provides acceptable validity within clinical recommendations for cup orientation, femoral offset and leg length in the supine patient position. With additional attention to the proposed mechanisms for error identified in the present analysis, the application of the imageless system of interest to pathologic anatomy in the context of challenging anthropometry, varying patient position and surgical approach, could become the standard for improving surgical outcomes in primary THA.

## Supporting information

Supplementary material 2 - Regression Models not used

Supplementary material 3 - Summary of validation findings

## Data Availability

Upon reasonable request to the authors, only de-identified data (i.e. data with sensitive and personal information removed), will be provided as per the ANDS Publishing and Sharing Sensitive Data Guide.

## Acknowledgements

The authors wish to acknowledge the important efforts of the staff of Sydney Bone and Joint Clinic, as well as Active Surgical (Ashley Pienaar and Francois Pienaar) in data collection for the study. In addition, the efforts of Milad Ebrahimi and Nalan Ektas (EBM Analytics) in assisting with data collection, data checking and analysis, and Manaal Fatima and Meredith Harrison-Brown (EBM Analytics) in assisting with the manuscript preparation are acknowledged.

## Supplementary material

Supplementary material 1 - Code

Supplementary material 2 - Regression model results summary

Supplementary material 3 - Summary of validation findings

