## Supplementary 1 - Code for "CT validation of intraoperative imageless navigation (Naviswiss) for component positioning accuracy in primary total hip arthroplasty in supine patient position: A prospective observational cohort study in a single-surgeon practice"

```
**Tidy and load
clear
cd "...\Statistical Analysis"
import delimited ".csv"
** Summary statistics for patient characteristics
summarize ageatsurgery bmi delta_inc_fpp delta_ver_fpp delta_total_offset delta_total_lld ns_inc ns_ver ns_offset ns_lld image_inc_fpp image_ver_fpp image_total_offset image_total_lld
*Fill missing BMI
summarize bmi
egen meanbmi = mean(bmi)
replace bmi = meanbmi if bmi ==.
****Part A - Calculate Agreement****
**Rock out with the bootstrap out - calculate median of delta
bootstrap r(p50), reps(1000) seed(1234) nodots: summarize delta_inc_app, detail
bootstrap r(p50), reps(1000) seed(1234) nodots: summarize delta_inc_fpp, detail
bootstrap r(p50), reps(1000) seed(1234) nodots: summarize delta_ver_app, detail
bootstrap r(p50), reps(1000) seed(1234) nodots: summarize delta_ver_fpp, detail
bootstrap r(p50) if !missing(delta_total_offset), reps(1000) seed(1234) nodots: summarize delta_total_offset, detail
bootstrap r(p50) if !missing(delta_total_lld), reps(1000) seed(1234) nodots: summarize delta_total_lld, detail
**Rock out with the bootstrap out - calculate mean of delta
bootstrap r(mean), reps(1000) seed(1234) nodots: summarize delta_inc_app, detail
bootstrap r(mean), reps(1000) seed(1234) nodots: summarize delta_inc_fpp, detail
bootstrap r(mean), reps(1000) seed(1234) nodots: summarize delta_ver_app, detail
bootstrap r(mean), reps(1000) seed(1234) nodots: summarize delta_ver_fpp, detail
bootstrap r(mean) if !missing(delta_total_offset), reps(1000) seed(1234) nodots: summarize delta_total_offset, detail
bootstrap r(mean) if !missing(delta_total_lld), reps(1000) seed(1234) nodots: summarize delta_total_lld, detail
**Rerun with absolute deviations (all declarations included)****
generate abs_delta_inc_app = abs(delta_inc_app)
generate abs_delta_inc_fpp = abs(delta_inc_fpp)
generate abs_delta_ver_app = abs(delta_ver_app)
generate abs_delta_ver_fpp = abs(delta_ver_fpp)
generate abs_delta_offset = abs(delta_total_offset)
generate abs_delta_lld = abs(delta_total_lld)
bootstrap r(mean), reps(1000) seed(1234) nodots: summarize abs_delta_inc_app, detail
bootstrap r(mean), reps(1000) seed(1234) nodots: summarize abs_delta_inc_fpp, detail
bootstrap r(mean), reps(1000) seed(1234) nodots: summarize abs_delta_ver_app, detail
bootstrap r(mean), reps(1000) seed(1234) nodots: summarize abs_delta_ver_fpp, detail
bootstrap r(mean) if !missing(abs_delta_offset), reps(1000) seed(1234) nodots: summarize abs_delta_offset, detail
bootstrap r(mean) if !missing(abs_delta_lld), reps(1000) seed(1234) nodots: summarize abs_delta_lld, detail
**Are the differences significantly different to Zero?
signrank delta_inc_app = 0
```

```
signrank delta_inc_fpp = 0
signrank delta_ver_app = 0
signrank delta_ver_fpp = 0
signrank delta_total_offset = 0
signrank delta_total_lld = 0

**Work out if ave is associated with delta_inc_app

bootstrap, reps(100) seed(123): regress delta_inc_fpp ave_inc_fpp
bootstrap, reps(100) seed(123): regress delta_inc_app ave_inc_app
bootstrap, reps(100) seed(123): regress delta_ver_fpp ave_ver_fpp
bootstrap, reps(100) seed(123): regress delta_ver_app ave_ver_app
bootstrap if !missing(delta_total_offset), reps(100) seed(123): regress delta_total_offset ave_total_offset
bootstrap if !missing(delta_total_lld), reps(100) seed(123): regress delta_total_lld ave_total_lld

cd "...\Statistical Analysis\ThirdPass\Original"

*Plot what would be Bland_Altman plots
*95%CI is prediction interval for individual datapoint (not the mean)
* line coordinates specified are retrieved from LOA calculations listed in [AnalysisTables] in NS Dashboard gsheet https://docs.google.com/spreadsheets/d/1InYb70d4dV-uehWHgJPTmLLKyltjMeRf8r_SzdXagFU/edit#gid=1365526240

twoway (lfitci delta_inc_fpp ave_inc_fpp, stdf acolor(gs6%50)) (scatter delta_inc_fpp ave_inc_fpp), yline(-10.7 6.9, lwidth(1pt) lcolor(blue) lpattern(dash))
graph export BlandAltmanInc.png, replace
graph close Graph
twoway (lfitci delta_ver_fpp ave_ver_fpp, stdf acolor(gs6%50)) (scatter delta_ver_fpp ave_ver_fpp), yline(-10.8 3.6, lwidth(1pt) lcolor(blue) lpattern(dash))
graph export BlandAltmanVer.png, replace
graph close Graph
twoway (lfitci delta_total_offset ave_total_offset, stdf acolor(gs6%50)) (scatter delta_total_offset ave_total_offset),yline(-8.2 11.2, lwidth(1pt) lcolor(blue) lpattern(dash))
graph export BlandAltmanOffset.png, replace
graph close Graph
twoway (lfitci delta_total_lld ave_total_lld, stdf acolor(gs6%50)) (scatter delta_total_lld ave_total_lld), yline(-6.6 10.8, lwidth(1pt) lcolor(blue) lpattern(dash))

graph export BlandAltmanLLD.png, replace
graph close Graph

cd "...\Statistical Analysis"

save "NS_JI Accuracy Analysis - Third Pass.dta", replace

***Part B factors associated with agreement: *No declarations omitted

* encode categorical variables

*rename gender sex
encode sex, gen(sexcode)

cd "...Statistical Analysis\ThirdPass\Original\Regression"

*Inclination
bootstrap, reps(100) seed(1234) nodots: regress delta_inc_fpp ns_inc c.bmi c.ageatsurgery i.sexcode, vce(robust)

*margins sexcode

predict delta_inc_predict, xb
```

```
generate new_ns_inc = ns_inc - delta_inc_predict
generate delta_inc_biasc = new_ns_inc - image_inc_fpp

generate abs_delta_inc_biasc = abs(delta_inc_biasc)

bootstrap r(mean), reps(1000) seed(1234) nodots: summarize delta_inc_biasc, detail
bootstrap r(mean), reps(1000) seed(1234) nodots: summarize abs_delta_inc_biasc, detail

twoway (lfitci delta_inc_biasc ave_inc_fpp, stdf acolor(gs6%50)) (scatter delta_inc_biasc ave_inc_fpp),yline(-7.2 7.2, lwidth(1pt) lcolor(blue) lpattern(dash))
graph export BlandAltmanIncBC.png, replace
graph close Graph

*Version
bootstrap, reps(100) seed(1234) nodots: regress delta_ver_fpp ns_ver c.bmi c.ageatsurgery i.sexcode, vce(robust)

predict delta_ver_predict, xb

generate new_ns_ver = ns_ver - delta_ver_predict
generate delta_ver_biasc = new_ns_ver - image_ver_fpp

generate abs_delta_ver_biasc = abs(delta_ver_biasc)

bootstrap r(mean), reps(1000) seed(1234) nodots: summarize delta_ver_biasc, detail
bootstrap r(mean), reps(1000) seed(1234) nodots: summarize abs_delta_ver_biasc, detail

twoway (lfitci delta_ver_biasc ave_inc_fpp, stdf acolor(gs6%50)) (scatter delta_ver_biasc ave_inc_fpp),yline(-6.7 6.7, lwidth(1pt) lcolor(blue) lpattern(dash))
graph export BlandAltmanVerBC.png, replace
graph close Graph

*Offset
bootstrap if !missing(delta_total_offset), reps(100) seed(1234) dots: regress delta_total_offset ns_offset c.bmi c.ageatsurgery i.sexcode, vce(robust)

predict delta_offset_predict, xb

*regress delta_offset_predict image_total_offset

generate new_ns_offset = ns_offset - delta_offset_predict
generate delta_offset_biasc = new_ns_offset- image_total_offset

generate abs_delta_offset_biasc = abs(delta_offset_biasc)

bootstrap r(mean) if !missing(delta_offset_biasc), reps(1000) seed(1234) nodots: summarize delta_offset_biasc, detail
bootstrap r(mean) if !missing(abs_delta_offset_biasc), reps(1000) seed(1234) nodots: summarize abs_delta_offset_biasc, detail

twoway (lfitci delta_offset_biasc ave_total_offset, stdf acolor(gs6%50)) (scatter delta_offset_biasc ave_total_offset),yline(-7.8 7.8, lwidth(1pt) lcolor(blue) lpattern(dash))
graph export BlandAltmanOffsetBC.png, replace
graph close Graph

*LLD
bootstrap if !missing(delta_total_lld), reps(100) seed(1234) dots: regress delta_total_lld ns_lld c.bmi c.ageatsurgery i.sexcode, vce(robust)

predict delta_lld_predict, xb
```

```
generate new_ns_lld = ns_lld - delta_lld_predict
generate delta_lld_biasc = new_ns_lld - image_total_lld

generate abs_delta_lld_biasc = abs(delta_lld_biasc)

bootstrap r(mean) if !missing(delta_lld_biasc), reps(1000) seed(1234) nodots: summarize delta_lld_biasc, detail
bootstrap r(mean) if !missing(abs_delta_lld_biasc), reps(1000) seed(1234) nodots: summarize abs_delta_lld_biasc, detail

twoway (lfitci delta_lld_biasc ave_total_lld, stdf acolor(gs6%50)) (scatter delta_lld_biasc ave_total_lld),yline(-8.6 8.6, lwidth(1pt) lcolor(blue) lpattern(dash)) ysc(r(-15 15))
graph export BlandAltmanlldBC.png, replace
graph close Graph

** Summary statistics for absolute deviations
summarize abs_delta_inc_app abs_delta_inc_fpp abs_delta_ver_app abs_delta_ver_fpp abs_delta_offset abs_delta_lld

**Rerun with absolute deviations (all declarations included)****

**Rock out with the bootstrap out - calculate mean of delta
bootstrap r(mean), reps(1000) seed(1234) nodots: summarize delta_inc_biasc, detail
bootstrap r(mean), reps(1000) seed(1234) nodots: summarize delta_ver_biasc, detail
bootstrap r(mean), reps(1000) seed(1234) nodots: summarize delta_offset_biasc, detail
bootstrap r(mean), reps(1000) seed(1234) nodots: summarize delta_lld_biasc, detail

bootstrap r(mean), reps(1000) seed(1234) nodots: summarize abs_delta_inc_biasc, detail
bootstrap r(mean), reps(1000) seed(1234) nodots: summarize abs_delta_ver_biasc, detail
bootstrap r(mean), reps(1000) seed(1234) nodots: summarize abs_delta_offset_biasc, detail
bootstrap r(mean), reps(1000) seed(1234) nodots: summarize abs_delta_lld_biasc, detail

*Declarations omitted
cd "...\Statistical Analysis\ThirdPass\DecOmit"

* encode intraop declarations


generate str3 intraopdec2 = ""
replace intraopdec2 = "No" if intraopdec == "Nothing to declare"
replace intraopdec2 = "Yes" if !(intraopdec == "Nothing to declare")
encode intraopdec2, gen(deccode)

**Run analysis with declarations removed

generate delta_offset_decomit =.
generate delta_lld_decomit =.
generate ageatsurgery_decomit =.
generate sexcode_decomit =.
generate bmi_decomit =.
generate ave_offset_decomit =.
generate ave_lld_decomit =.
generate ns_offset_decomit =.
generate ns_lld_decomit =.

replace delta_offset_decomit = delta_total_offset if deccode ==1
replace delta_lld_decomit = delta_total_lld if deccode ==1
replace ageatsurgery_decomit = ageatsurgery if deccode ==1
replace sexcode_decomit = sexcode if deccode ==1
replace bmi_decomit = bmi if deccode ==1
replace ave_offset_decomit = ave_total_offset if deccode ==1
replace ave_lld_decomit = ave_total_lld if deccode ==1
replace ns_offset_decomit = ns_offset if deccode ==1
```

```
replace ns_lld_decomit = ns_lld if deccode ==1

**Rock out with the bootstrap out - calculate mean of delta
bootstrap r(mean) if !missing(delta_offset_decomit), reps(1000) seed(1234) nodots: summarize
delta_offset_decomit, detail
bootstrap r(mean) if !missing(delta_lld_decomit), reps(1000) seed(1234) nodots: summarize
delta_lld_decomit, detail

**Rerun with absolute deviations

generate abs_delta_offset_decomit = abs(delta_offset_decomit)
generate abs_delta_lld_decomit = abs(delta_lld_decomit)


*Plot Bland-Altman
twoway (lfitci delta_offset_decomit ave_offset_decomit, stdf acolor(gs6%50)) (scatter
delta_offset_decomit ave_offset_decomit),yline(-8.8 11.4, lwidth(1pt) lcolor(blue) lpattern(dash))
graph export BlandAltmanOffsetDecOmit.png, replace
graph close Graph
twoway (lfitci delta_lld_decomit ave_lld_decomit, stdf acolor(gs6%50)) (scatter delta_lld_decomit
ave_lld_decomit),yline(-6.5 11.3, lwidth(1pt) lcolor(blue) lpattern(dash))
graph export BlandAltmanLLDDecOmit.png, replace
graph close Graph

**Rock out with the bootstrap out - calculate mean of delta
bootstrap r(mean) if !missing(delta_offset_decomit), reps(1000) seed(1234) nodots: summarize
delta_offset_decomit, detail
bootstrap r(mean) if !missing(delta_lld_decomit), reps(1000) seed(1234) nodots: summarize
delta_lld_decomit, detail

bootstrap r(mean) if !missing(delta_offset_decomit), reps(1000) seed(1234) nodots: summarize
abs_delta_offset_decomit, detail
bootstrap r(mean) if !missing(delta_lld_decomit), reps(1000) seed(1234) nodots: summarize
abs_delta_lld_decomit, detail

*Maybe only need up to here for now 12-Nov-2022


cd "...\Statistical Analysis"


**Clean up and close


save "...Analysis - Third Pass.dta", replace

export delimited "... - Output"

clear
```
