## Supplementary material 2 - Regression Models not used for "CT validation of intraoperative imageless navigation (Naviswiss) for component positioning accuracy in primary total hip arthroplasty in supine patient position: A prospective observational cohort study in a single-surgeon practice"

Table 1: Regression summary delta inclination (FPP)

```
Linear regression                              Number of obs =     34
                                               Replications  =    100
                                               Wald chi2(4)  =   7.25
                                               Prob > chi2   = 0.1233
                                               R-squared     = 0.2994
                                               Adj R-squared = 0.2028
                                               Root MSE      = 3.9841
```

| delta_in~fpp | Observed coefficient | Bootstrap std. err. | z | P>\|z\| | Normal-based [95% conf. | interval] |
|---|---|---|---|---|---|---|
| ns_inc | -.7202958 | .2922558 | -2.46 | 0.014 | -1.293107 | -.147485 |
| bmi | -.2253103 | .1315003 | -1.71 | 0.087 | -.4830463 | .0324256 |
| ageatsurgery | -.0356231 | .0797826 | -0.45 | 0.655 | -.1919941 | .1207479 |
| sexcode | | | | | | |
| Male | .8781924 | 1.416129 | 0.62 | 0.535 | -1.89737 | 3.653755 |
| _cons | 36.44397 | 15.52767 | 2.35 | 0.019 | 6.01029 | 66.87765 |

Table 2: Regression summary delta version (FPP)

```
Linear regression                              Number of obs =      34
                                               Replications  =     100
                                               Wald chi2(4)  =    4.68
                                               Prob > chi2   =  0.3222
                                               R-squared     =  0.1154
                                               Adj R-squared = -0.0066
                                               Root MSE      =  3.7988
```

| delta_ve~fpp | Observed coefficient | Bootstrap std. err. | z | P>\|z\| | Normal-based [95% conf. | interval] |
|---|---|---|---|---|---|---|
| ns_ver | -.2725811 | .1661733 | -1.64 | 0.101 | -.5982748 | .0531126 |
| bmi | -.1300883 | .1714082 | -0.76 | 0.448 | -.4660422 | .2058656 |
| ageatsurgery | .0334747 | .0745442 | 0.45 | 0.653 | -.1126293 | .1795787 |
| sexcode | | | | | | |
| Male | 1.628467 | 1.44538 | 1.13 | 0.260 | -1.204427 | 4.461361 |
| _cons | 2.665045 | 7.352848 | 0.36 | 0.717 | -11.74627 | 17.07636 |

Table 3: Regression summary delta total offset

```
Linear regression                               Number of obs =         32
                                                Replications  =        100
                                                Wald chi2(4)  =      13.56
                                                Prob > chi2   =     0.0088
                                                R-squared     =     0.3512
                                                Adj R-squared =     0.2550
                                                Root MSE      =     4.1696
```

| delta_tota~t | Observed coefficient | Bootstrap std. err. | z | P>\|z\| | Normal-based [95% conf. | interval] |
|---|---|---|---|---|---|---|
| ns_offset | .569177 | .1869792 | 3.04 | 0.002 | .2027044 | .9356496 |
| bmi | .0089397 | .2717262 | 0.03 | 0.974 | -.5236338 | .5415132 |
| ageatsurgery | -.0563344 | .0954872 | -0.59 | 0.555 | -.2434859 | .130817 |
| sexcode | | | | | | |
| Male | .865698 | 2.410951 | 0.36 | 0.720 | -3.859678 | 5.591074 |
| _cons | 5.854657 | 12.20774 | 0.48 | 0.632 | -18.07207 | 29.78138 |

Table 4: Regression summary delta leg length difference

```
Linear regression                               Number of obs =         32
                                                Replications  =        100
                                                Wald chi2(4)  =       0.84
                                                Prob > chi2   =     0.9328
                                                R-squared     =     0.0439
                                                Adj R-squared =    -0.0977
                                                Root MSE      =     4.6307
```

| delta_tota~d | Observed coefficient | Bootstrap std. err. | z | P>\|z\| | Normal-based [95% conf. | interval] |
|---|---|---|---|---|---|---|
| ns_lld | .1194275 | .2612432 | 0.46 | 0.648 | -.3925997 | .6314547 |
| bmi | -.1118321 | .2165626 | -0.52 | 0.606 | -.536287 | .3126228 |
| ageatsurgery | .0281283 | .1291462 | 0.22 | 0.828 | -.2249936 | .2812501 |
| sexcode | | | | | | |
| Male | 1.293895 | 1.850367 | 0.70 | 0.484 | -2.332758 | 4.920548 |
| _cons | 1.841925 | 11.61245 | 0.16 | 0.874 | -20.91806 | 24.60191 |
