## Supplementary material 3 - Summary of validation findings for "CT validation of intraoperative imageless navigation (Naviswiss) for component positioning accuracy in primary total hip arthroplasty in supine patient position: A prospective observational cohort study in a single-surgeon practice"

| | This study (sign) | This study (absolute) | Liu et al 2023 (sign) | Liu et al 2023 (absolute) | Hasegawa et al 2022[1] (supine - absolute) | Pooled[1–8] (N = 688) (absolute) |
|---|---|---|---|---|---|---|
| Inclination_ FPP (°) | -1.9 (4.5) | 3.3 (3.6) | 1.0 (4.6) | 3.6 (3.1) | | |
| Inclination (Bias Corrected) | -0.002 (3.7) | 3 (2.1) | 0 (4.0) | 3.2 (2.7) | 2.8 (2.2) | 2.8 (2.0) |
| Version_ FPP (°) | -3.6 (3.7) | 4.3 (3.0) | 2.0 (4.5) | 4.0 (2.6) | | |
| Version (Bias Corrected) | 0 (3.4) | 2.8 (2.2) | 0 (4.0) | 3.4 (2.2) | 2.8 (2.0) | 3.6 (3.5) |
| Offset (mm)* | 1.3 (5.1) | 3.9 (3.3) | 2.1 (2.4) | 2.4 (2.1) | - | - |
| LLD (mm)* | 2.4 (4.5) | 3.7 (3.2) | 0.4 (2.4) | 1.8 (1.3) | - | - |

*Declarations omitted
